# Abusers indoors and coronavirus outside: an examination of public discourse about COVID-19 and family violence on Twitter using machine learning

**DOI:** 10.1101/2020.08.13.20167452

**Authors:** Jia Xue, Junxiang Chen, Chen Chen, Ran Hu, Tingshao Zhu

**Author notes:** **Corresponding author**: Jia Xue, Assistant Professor, Factor-Inwentash Faculty of Social Work & Faculty of Information, University of Toronto, 246 bloor street west, Toronto, ON, Canada.

## Abstract

**Purpose:** This brief report aims to provide the first large-scale analysis of public discourse regarding family violence and the COVID-19 pandemic on Twitter. Method: We analyzed 301,606 Tweets related to family violence and COVID-19 from April 12 to July 16, 2020, for this study. We used the machine learning approach, Latent Dirichlet Allocation, and identified salient themes, topics, and representative Twitter examples.

**Results:** We extracted nine themes on family violence and COVID-19 pandemic, including (1) the Impact of COVID-19 on family violence (e.g., rising rates, hotline calls increased, murder & homicide); (2) the types (e.g., child abuse, domestic violence, sexual violence) and (3) forms of family violence (e.g., physical aggression, coercive control); (4) risk factors of family violence (e.g., alcohol abuse, financial constraints, gun, quarantine); (5) victims of family violence (e.g., LGBTQ, women, and women of color, children); (6) social services of family violence (e.g., hotlines, social workers, confidential services, shelters, funding); (7) law enforcement response (e.g., 911 calls, police arrest, protective orders, abuse reports); (8) Social movement/awareness (e.g., support victims, raise awareness); and (9) domestic violence-related news (e.g., Tara Reade, Melissa Derosa).

**Conclusions:** The COVID-19 has an impact on family violence. This report overcomes the limitation of existing scholarship that lacks data for consequences of COVID-19 on family violence. We contribute to the understanding of family violence during the pandemic by providing surveillance in Tweets, which is essential to identify potentially effective policy programs in offering targeted support for victims and survivors and preparing for the next wave.

## Introduction

The previous study indicates that epidemic disease (e.g., Eloba) increases the rates of domestic violence (International Rescue Committee, 2019). Rigorous measures (e.g., quarantine, social isolation) are effective to control the spread of COVID-19, but they bring a series of negative social consequence, such as negative psychological stresses (Galea, Merchant & Lurie, 2020; Li et al., 2020; Su et al., 2020), growth of unemployment (Roje Ðapić et al., 2020), and increased rates of violence against women and children (Bradbury-Jones & Isham, 2020; Campbell, 2020; Peterman et al., 2020; WHO, 2020). In many countries, the reported cases and service needs of family violence dramatically increase since the quarantine measures came into force (Salisbury, 2020). In the UK, calls to Domestic Violence Helpline increase by 25% in the first week after the lockdown (BBC, 2020). Domestic violence cases increase three times in Hubei province in China during the lockdown (Fraser, 2020). In the United States, there is a 10.2% increase in domestic violence calls during the COVID-19 pandemic (Leslie and Wilson, 2020). These reports illustrate that existing COVID-19 intervention measures may be having a profound impact on family violence victims. However, there is a lack of data on COVID-19 pandemic and family violence (Anurudran et al., 2020), which demonstrates further research. During the implementation of the social isolation measures, social media should be leveraged to raise public awareness and share best practices (e.g., “bystander approaches,” “supportive statements,” “accessing help on behalf of a survivor”) (Van Gelder et al., 2020, p.2), and provide support (Boserup, McKenney & Elkbuli, 2020). Twitter has been used to examine the nature of domestic violence (Cravens, Whiting & Aamar, 2015; Xue et al., 2019a, b). For example, a significant of studies describe Twitter hashtag#MeToo as a phenomenon tool for disclosing sexual harassment experience, and more importantly, ignite a widespread social campaign or political protest.

This brief report aims to provide the first large-scale analysis of public discourse regarding family violence on Twitter using machine learning techniques during the COVID-19 pandemic. The research questions include (1) What contents are discussed relating to family violence and COVID-19? and (2) what themes are identified relating to family violence and COVID-19? An examination of Twitter data related to family violence and COVID-19 offers a new perspective to capture and understand the impact and the risk factors associated with COVID-19 on family violence, which is essential for developing policy programs in responding and mitigating the adverse effects and offering targeted support for victims and survivors (Chandan et al., 2020).

## Methods

### Data collection and sample

The sampling frame was our COVID-19 dataset between April 12 to July 16, 2020, which used a list of COVID-19 relevant hashtags as search terms to randomly collect Tweets from Twitter (Xue et al., 2020a, b). Figure 1 showed the Tweets pre-processing chart. Our dataset included a total of 274,501,992 Tweets during this period, within which 186,678,079 were English Tweets, and 87,823,913 were non-English Tweets. We sampled Tweets using keywords, including “domestic violence,” “intimate partner violence,” “family violence,” “violence against women,” “gender-based violence,” “child abuse,” “child maltreatment,” “elder abuse” and “IPV.” We fetched 1,015,874 Tweets from this dataset. After removing 714,268 duplicates, we analyzed a total of 301,606 remaining Tweets for this study.

**Figure 1.**
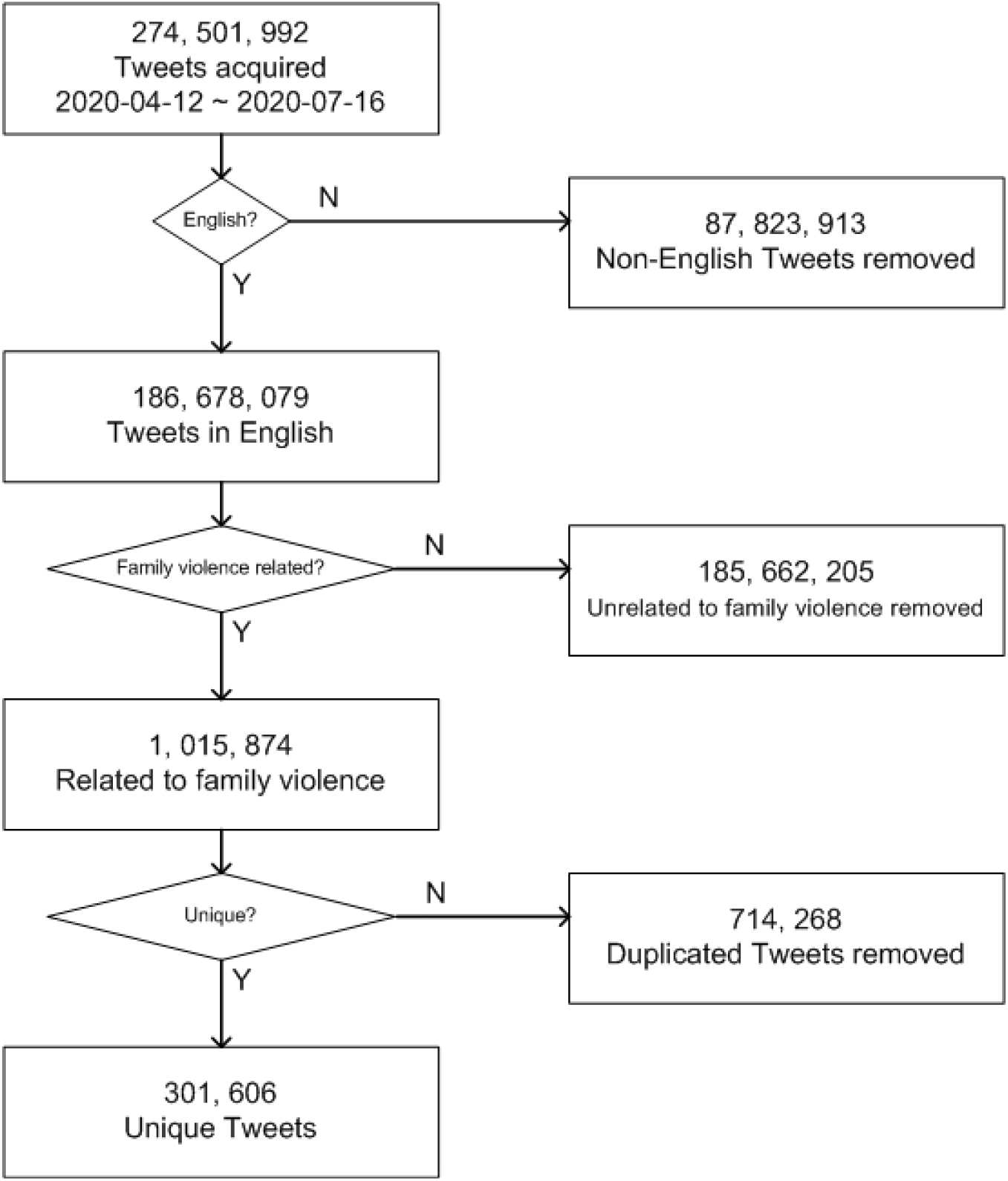
Tweets pre-processing chart.

### Data analysis

**Pre-processing the raw data**. We used Python to clean the data and removed the following items because they had no contribution to the semantic meanings of the Tweets, including the hashtag symbol, URLs, @users, special characters, punctuations, and stop-words.

**Unsupervised machine learning**. We used a machine learning approach, Latent Dirichlet Allocation (LDA, Blei et al., 2003), to analyze a corpus of unstructured text. LDA was a generative statistical model that regards a corpus of text (Tweets) as a mixture of a small number of latent topics. Each latent topic was assigned with a set of linguistic units (e.g., single words or a pair of words) that were counted by the algorithm. These linguistic units with high frequency were likely to co-occur together and form into different latent topics. With the LDA model, the distribution of topics in documents can be inferred. LDA assumed a generative process describing how the documents were created, such that we can infer or reverse engineer the topic distributions. The generative process of LDA for M documents, each of which had the length of N_i_ was given as:

1. Choose *θ_i_* ~ Dir(*α*), with *i* ∊ {1,…*,M*}.
2. Choose *ϕ_k_* ~ Dir(*β*), with *k* ∊ {1,…*,K*}.
3. For the j-th linguistic unit in the i-th document with *i* ∊ {1,…, *M}*, and *j* ∊ {1,…, *N_i_*},

a. Choose *z_ij_* ~ Multinomial(*θ_i_*).
b. Choose *w_i,j_* ~ Multmomial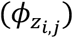.

Table 1 showed the meaning of the notations. With the generative process described above, the distributions of the topics can be inferred using the *python* package *genism*.

**Table 1.**
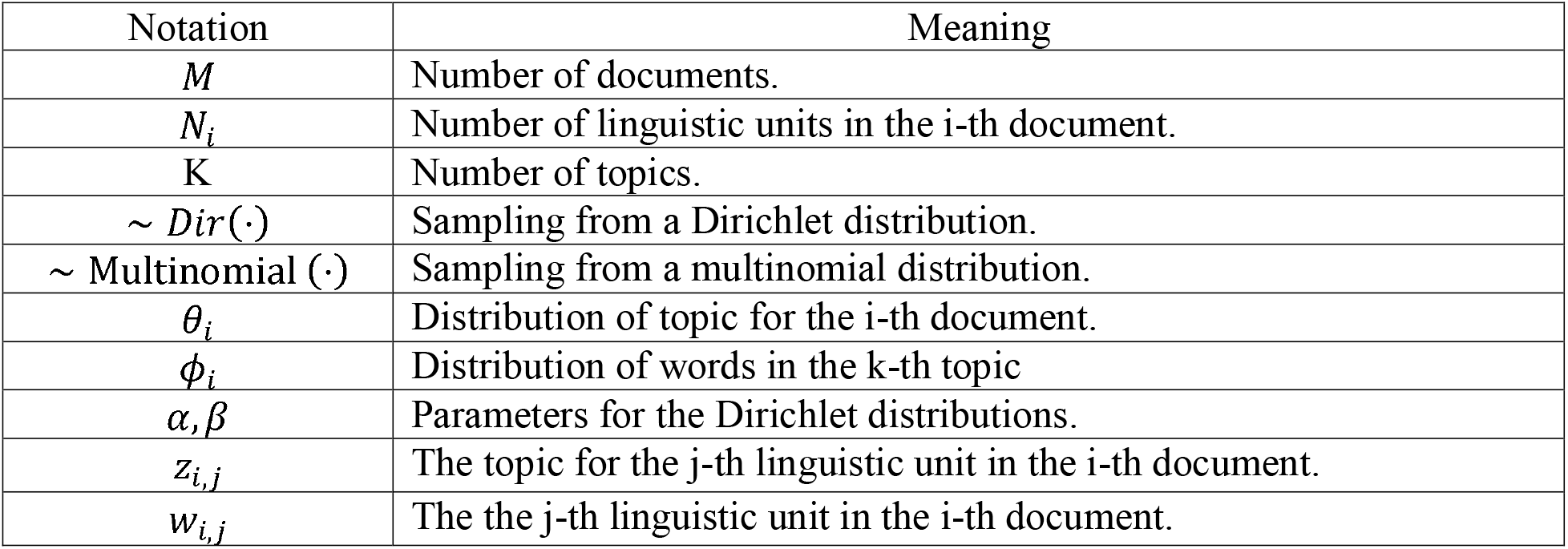
Notations for Latent Dirichlet Allocation (LDA)

## Results

Between April 12 to July 16, 2020, we analyzed 301,606 Tweets that discussed family violence and COVID-19. We identified 50 latent topics and the frequently mentioned pairs of words under each topic. We further categorized these 50 identified common topics into 9 themes and 30 categories, shown in Table 2. Table 2 showed commonly co-occurring bigrams (pair of words) and representative Tweets examples under each identified theme and categories.

**Table 2.**
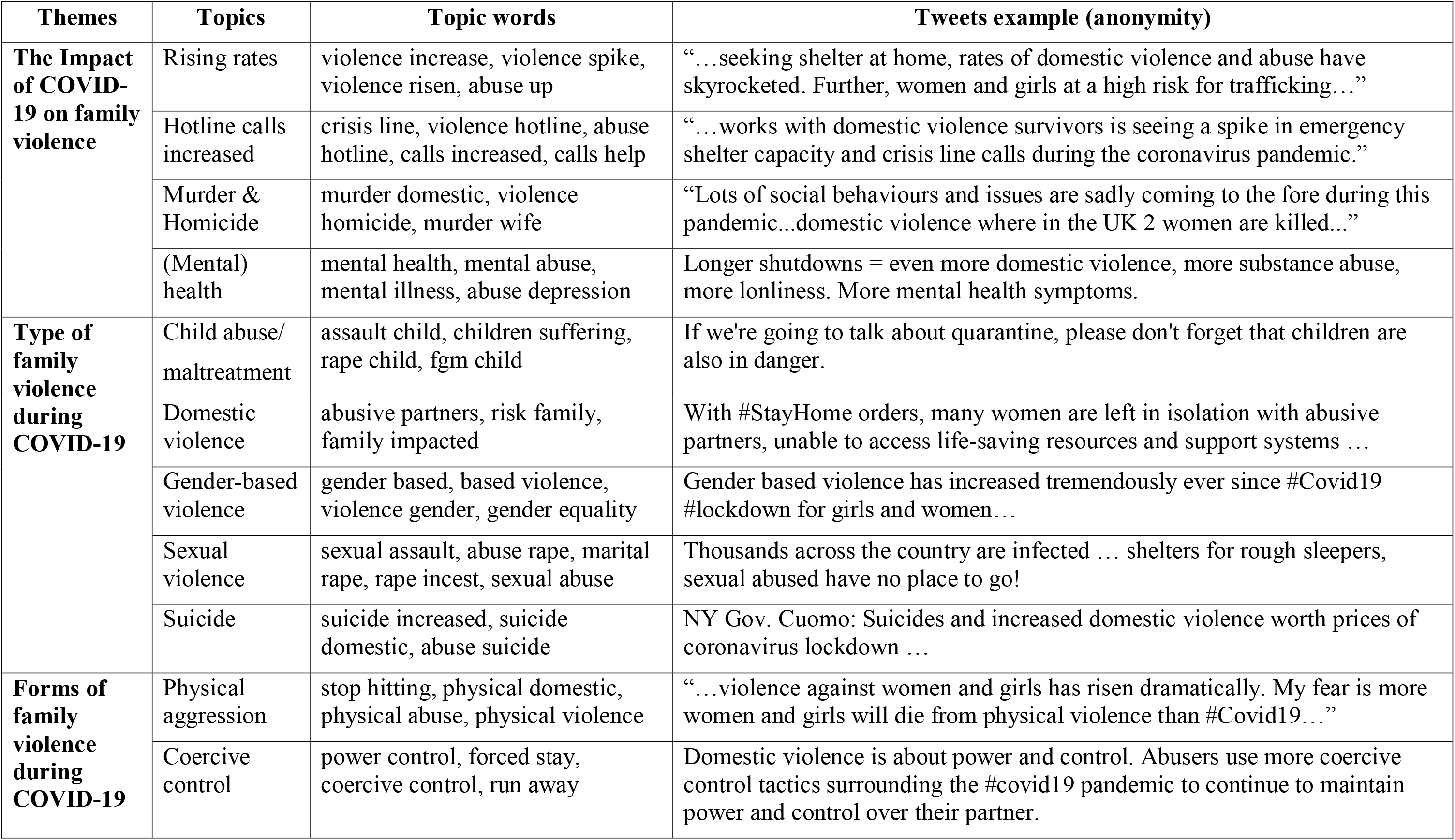
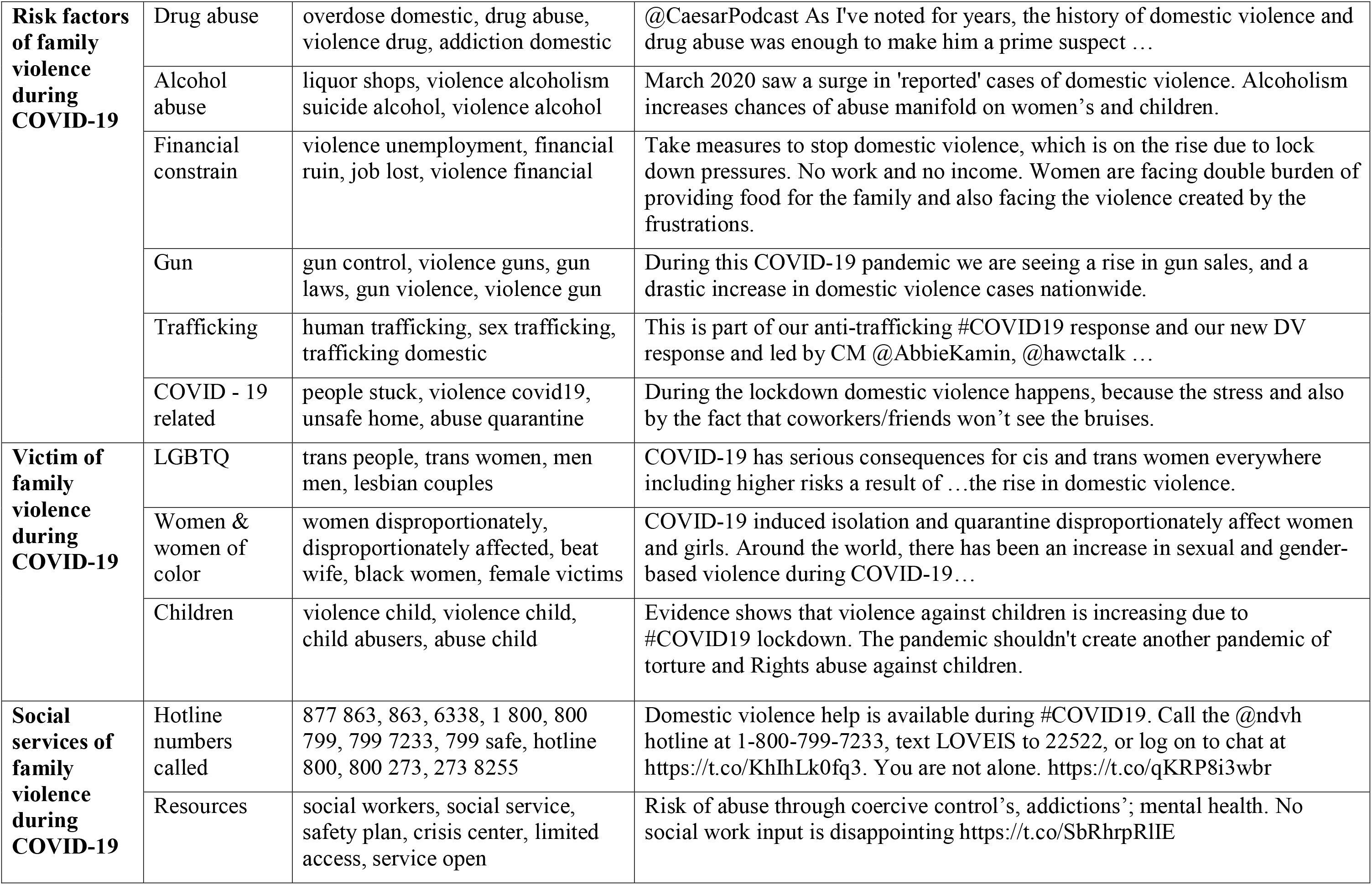
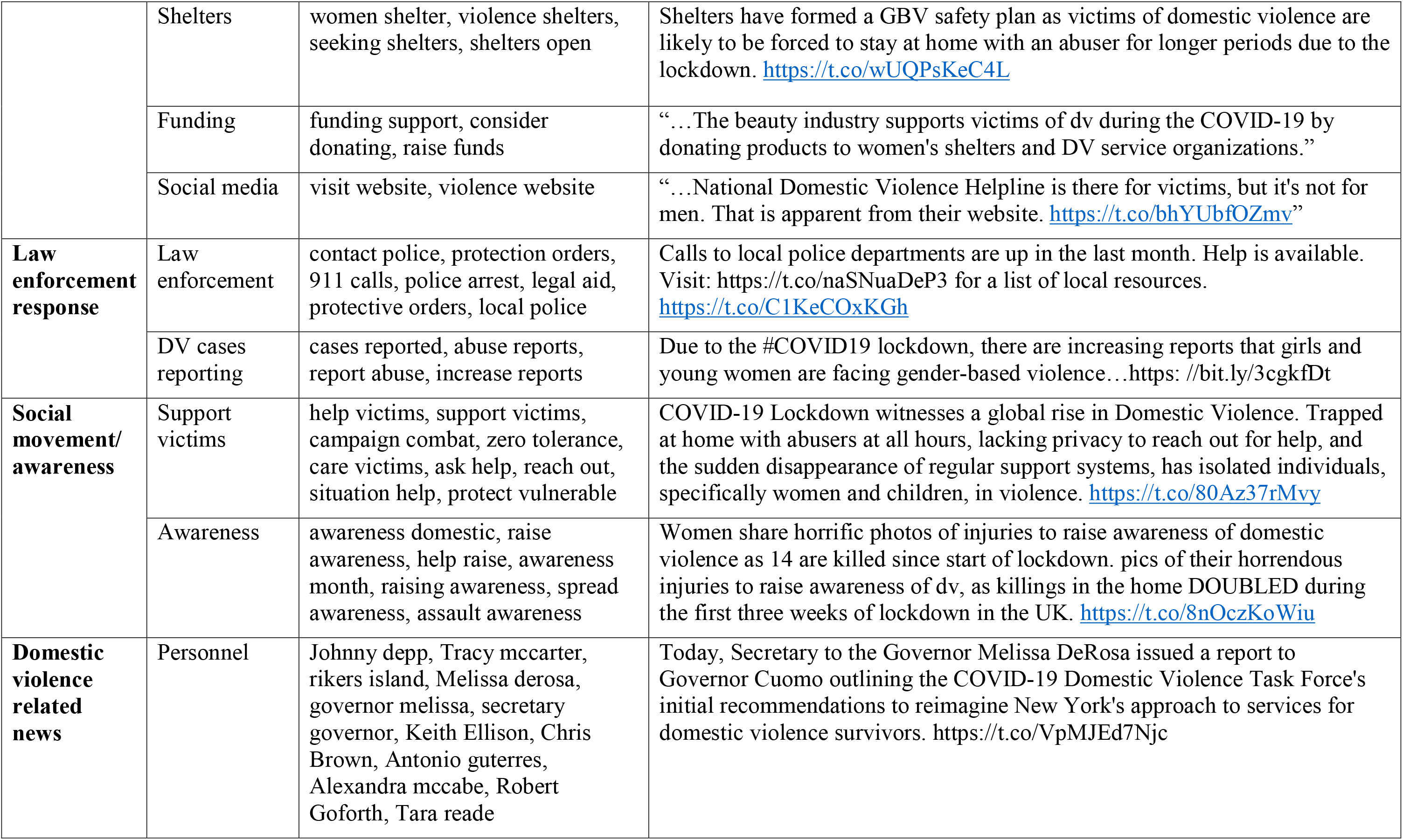
Themes, topics, commonly co-occurred topics, and representative tweets examples from domestic violence and COVID-19 related Tweets.

***Theme 1: The Impact of COVID-19 on family violence***. The high frequency of domestic violence rising rates-related Tweets indicated that COVID-19 was having an impact on family violence, including popular bigrams “violence increased,” “violence higher,” “rising violence,” and “violence skyrocketing.” Increased Hotline calls was another impact of COVID-19 on family violence (e.g., calls increased, calls help). As a representative Tweet message indicated, “A Miami Valley nonprofit agent is seeing a spike in crisis line calls during the pandemic.” In addition, the pandemic had consequences, including domestic violence murder and homicide, and mental health (e.g., depression, mental abuse) issues.

***Theme 2: Type of family violence during COVID-19***. Findings showed that several types of family violence were affected during COVID-19, including “child abuse/maltreatment (e.g., assault child, rape child),” “domestic violence (e.g., abusive partners, violence partners),” “gender-based violence (e.g., gender-based violence, gender equality),” “sexual violence (e.g., sexually assault, marital rape),” and “suicide (e.g., suicide increased, suicide domestic).”

***Theme 3: Forms of family violence during COVID-19***. Two primary forms of family violence were discussed on Twitter during the COVID-19, including “physical aggression (e.g., physically hurt, stop hitting),” and “coercive control (e.g., power control, forced stay).” For example, “…abusers may use more coercive control tactics surrounding the #covid19 pandemic to continue to maintain power and control over their partner.”

***Theme 4: Risk factors of family violence during COVID-19***. We found that the rising rate of domestic violence was associated with risk factors, including “drug abuse,” “alcohol abuse,” “financial constraints (e.g., job loss, loss income),” “gun,” “trafficking,” and “COVID-19 related (e.g., lockdown, stuck home, quarantine, stuck home).” For example, “March 2020 saw a surge in reported cases of domestic violence. Alcoholism increases chances of abuse manifold on women and children…” and “During the lockdown, domestic violence happens because the coworkers/friends can’t see the bruises.”

***Theme 5: Victim of family violence during COVID-19***. LGBTQ community, women, women of color, and children consisted of the family violence victims during the COVID-19, indicated by the Tweets. Popular words in describing the victims and survivors of family violence included “trans people,” “lesbian couples,” “women disproportionately affected,” “beat wife,” “black women,” “female victims,” “violence child,” “child abusers,” and “abuse child.”

***Theme 6: Social services of family violence during COVID-19***. Social services of family violence were a main theme discussed by Twitter users during the COVID-19, indicated by the high-frequency mentions of hotline numbers. Resources, shelters, funding support, and visiting family violence websites were also frequently mentioned social services in the Tweets. In addition, social workers, confidential services, safety plans, and limited access were representative topics identified in the Tweets.

***Theme 7: Law enforcement response***. With the rising rates of family violence during the pandemic, DV cases reporting (e.g., “cases reported,” “abuse reports,” “violence reports,” “increase reports,” and “reported increase.”) was identified as a salient topic in the Tweets. Police departments (e.g., “police officers,” “local police,” “police chief,” “911 calls,” “contact police,” “police arrest”) were the first responders on the front lines of increased domestic violence reports during the COVID-19.

***Theme 8: Social movement/ awareness***. Findings also identified a social justice movement/awareness to support family violence victims and survivors. Tweets contents highlighted the advocacy and zero tolerance of the domestic violence, indicated by popular pair of words such as “help victims,” “campaign combat,” “violence advocacy,” “care victims,” “raise awareness,” and “awareness campaign.” For example, “Women share horrific photos of injuries to raise awareness of domestic violence...”

***Theme 9: Domestic violence-related news***. News events related to domestic violence cases during the COVID-19 were also identified, such as (1) the American actor Johnny Depp’s denied the domestic abuse allegations to his ex-wife Amber Heard; (2) Tracy McCarter was accused of stabbing her husband to death in Manhattan; (3) the singer Chris Brown arrested in Paris on allegations of rape; (4) Tara Reade’ sexual assault allegations by Joe Biden; and (5) Kentucky legislator Robert Goforth arrested on fourth-degree domestic violence.

News of solutions to help domestic violence survivors were also frequently discussed in the Tweets from April to July. For example, the governor of the New York State Council on Women and Girls, Melissa DeRosaand, created a task force to find innovative solutions to the domestic violence spike during the COVID-19 pandemic. The UN chief Antonio Guterres called for measures to address the surge in domestic violence linked to lockdowns that were imposed by governments in responding to the COVID-19 pandemic.

## Discussion

### Principal results

This brief report provides the first large-scale analysis of Tweets regarding public discourse of family violence and COVID-19. Twitter data in the study consists of a random of 301,606 Tweets relating to both family violence and coronavirus from April 12 to July 16, 2020. Machine learning technique LDA extracts high frequency of co-occurred pairs of words, and topics related to family violence among large-scale unstructured Tweets. The study contributes to the literature on family violence during the COVID-19 pandemic, including (1) the Impact of COVID-19 on family violence (e.g., increasing rates, victims affected); (2) the types and (3) forms of family violence during COVID-19; (4) victims of family violence; (5) risk factors of family violence during COVID-19; (6) social services of family violence during COVID-19; (7) law enforcement response; (8) Social movement/ awareness; and (9) domestic violence-related news. The present study overcomes the limitation of existing scholarship that lacks data or anecdotal reports. We contribute to the understanding of family violence during the pandemic by providing surveillance in Tweets, which is essential to identify potentially effective policy programs in offering targeted support for victims and survivors and preparing for the next wave.

### Victims of family violence and the COVID-19 pandemic

Our results provide insights about who are at higher risks of family violence during the lockdown. Findings reveal a broader range of family violence regardless of gender, such as the LGBTQ community (e.g., men men). To compare our results with one recent study using Twitter data for domestic violence research (Xue et al., 2019a), we also find that domestic violence-related discussions focus on the support and protection of victims instead of interventions against abusers. Women and children are disproportionately affected by family violence that is consistent with the majority of the research in the field (Bradbury-Jones & Isham, 2020; Fantuzzo et al., 1997; Tjaden & Thoennes, 1998; Xue, Chen, & Gelles, 2019a). Violence against children is associated with the characteristics of a disaster in previous epidemic studies (Roje Ðapić et al., 2020). UNICEF (2020) reports that school closures contribute to the increasing rates of child (sexual) abuse and neglect during the Ebola epidemic. It is also important to note that child abuse and domestic violence are likely to co-occur when isolated at home (Humphreys, Myint & Zeanah, 2020; Vu, Jouriles, McDonald & Rosenfield, 2016). In addition, our results provide evidence to support that Twitter has a limitation in primarily posting stories about male-to-female violence (Cravens, Whiting & Aamar, 2015) even though other patterns of violence exits, including female-to-male, male-to-male and bidirectional intimate partner violence (Hu, Xue, Lin, Sun & Wang, 2019).

### Risk factors of family violence in responding to the COVID-19

Our findings identify that a range of risk factors are associated with family violence during pandemics, such as drug abuse, alcohol abuse, financial constraints, gun, trafficking, and COVID-19 lockdown and social distancing. Our study reveals similar results with one recent report by Peterman and colleagues (2020) who summarize that nine main pathways are connecting the COVID-19 pandemic and violence against women and children, such as “economic insecurity and poverty-related stress (p.6),” “quarantines and social isolation (p.9),” “disaster and conflict-related unrest and instability (p.10),” and “inability to temporarily escape abusive partners (p.15).” Alcohol abuse continues to be a risk factor for family violence during stressful events (Catala-Minana et al., 2017). Financial constraints (e.g., financial ruin, lost jobs, economic collapse) linking COVID-19 create a barrier for family violence victims when they seek help (Van Gelder et al., 2020). Beland and colleges (2020) analyze the Canadian perspective survey and find that financial worries due to COVID-19 contribute to the increase of family violence and stress. An increasing rate of domestic homicides identified in Tweets suggests that guns are still a concern at home where family violence occurs.

Specific COVID-19 related risk factors (e.g., quarantines, social isolation) limit victims contact with the outside world, which are indicated by frequent use of words, such as “people stuck,” “unsafe home,” “people locked,” and “abuse quarantine” on Twitter. Since the rigorous measures overlap with many of the intervention strategies for family violence (Van Gelder et al., 2020), they are likely to increase the vulnerability of victims, such as increased exposure to the exploitative relationship, reduced options for support (Peterman et al., 2020), economic stress (US Bureau of Labor Statistics, 2020) and alcohol abuse (Colbert et al., 2020; Malathesh, Das & Chatterjee, 2020). For example, isolation limits the social contact with families and social services and thus facilitates violence and prevent victims from seeking help (Gelles, 1983; Gelles and Straus, 1979; Hagan, Raghavan & Doychak, 2019). During the COVID-19, home becomes a very dangerous place for victims, while individuals are living in forced close quarters (Salisbury, 2020). Besides, mental health exacerbated by social isolation increases the likelihood of locking domestic violence victims in an unsafe home environment and increases their vulnerability (Khalifeh, 2010).

### Social support system addressing family violence

Multi-agency integrating the law enforcement responses (e.g., protection orders, arrest), social services (e.g., hotlines, shelters), and social awareness/movements are recommended to address domestic violence and support victims (Gulati & Kelly, 2020). It is vital that social services (including social work practitioners, therapists) for domestic violence and sexual violence are resourced during the pandemic. Due to the mobility restriction, a lack of informal support, such as family, friends, co-workers, further contributes to the increased rates of family violence during the pandemic. Thus, it is more crucial than ever that victims can access support from the voluntary sector practitioners during the COVID-19 pandemic (Bradbury-Jones & Isham, 2020). Our results provide evidence that some agencies continue their roles during the pandemic. For example, several hotline numbers in the US have been frequently mentioned crisis lines during the pandemic, such as “Illinois Domestic Violence Hotline, 877-863-6338 (877-TO END DV),” “National Suicide Prevention Lifeline, 800-273-8255 (US),” “National Domestic Violence Hotline, 800-799-SAFE (7233) (US),” “National Sexual Assault Telephone Hotline, 800-656-HOPE (4673) (US),” and “Loveisrespect, Text LOVEIS to 22522 (US).” We also identified popular hotline numbers from the UK, such as Helplines/mind the mental health (Infoline: 0300-123-3393), national stalking helpline (0808-802-0300), and national domestic abuse helpline (0808-2000-247). However, a commentary in the *Canadian Medical Association Journal* raises concerns about the family violence support in the videoconference or telemedicine settings where the abusers are present (Bradley et al., 2020). Abusers can coercively control victims-survivors to use mobile phones for hotline support. Therefore, further evidence is needed to indicate whether the services fulfill their roles.

### Publicized cases of family violence on Twitter

Twitter conversations about highly publicized domestic violence cases are significant. News about the Hollywood star Johnny Depp denied his abuse allegations to the British judge when he was accused of domestic violence against his ex-wife Amber Heard is a prominent topic in our Tweets. Our results show public discussions of high-profile cases of domestic violence (e.g., sports-related domestic violence figures arrested for domestic violence), which is consistent with previous studies. Cravens, Whiting, and Aamar (2015) use qualitative content analysis to examine the factors that influence victims of IPV to leave an abusive relationship using 676 Tweets related to #whyIstayed and #whyIleft. Xue, Chen, and Gelles (2019a) analyze 322,863 Tweets about domestic violence and find that high-profile cases are prominent such as Greg Hardy’s domestic violence case. These studies consistently show that Twitter continues to be a source of news coverage on current events for domestic violence, even during the COVID-19 pandemic.

### Limitations

There are a couple of limitations. First, Twitter data has a limitation in collecting insights from online Twitter users, and thus do not represent the opinions of the whole population. Despite this shortcoming, this study provides the first large-scale analysis using Tweets, which allows researchers to identify the real-time data on the impact of COVID-19 on family violence. Second, the study does not include non-English Tweets in the analysis. Future studies could expand the literature by contributing to the findings from non-English Tweets analysis regarding the impact of COVID-19 on family violence. Third, even though our collected data covers 90 days of the outbreak since April 12, 2020, discussion patterns may continue to evolve with the changes of the COVID-19 situation over time. Fourth, to protect the privacy and anonymity of Twitter users, the present study does not examine the socio-demographic characteristics of the sample. It remains unknown whether the collected Tweets were from victims, perpetrators, organizations.

## Conclusion

COVID-19 has an impact on family violence, which is identified in a large-scale Twitter dataset. Our results reveal 9 themes and 30 topics relating to family violence and the COVID-19 pandemic. This brief report’s findings have implications for research that Twitter can provide a platform for real-time and large-scale surveillance of family violence by offering an understanding of the impact and risk factors associated with COVID-19, which is essential for developing policy programs for supporting victims and survivors. Findings provide insights for professionals who work with family violence victims and survivors regarding how to develop a social network-based support system for both informal and formal help when the conventional in-person support is unavailable during the second wave.

## Data Availability

This study followed Twitters Developers terms and policy (https://developer.twitter.com/en/developer-terms/policy) which has restrictions on redistributing Twitter content to third parties. We have fetched Twitter data between April 11 to July 16 2020 using a list of COVID-19 related hashtags as search terms and family-related terms as screen terms. We provided sufficient information about these hashtags in the paper. Other researchers can purchase a similar dataset during this period via Twitters service https://support.gnip.com/apis/historical_api2.0/

## Acknowledgements

The data collection is supported by Artificial Intelligence Lab for Justice at University of Toronto, Canada. http://aij.utoronto.ca

## Notes

### Competing Interest Statement

The authors have declared no competing interest.

### Funding Statement

Op Grant: COVID-19 Rapid Research FO - Social Policy and Public Health Responses

### Author Declarations

We used public Twitter data and exempted from IRB

